# Decision Support in Publicly Available Patient Information Policies at U.S. Osteopathic Medical Schools: A Vignette-Based Document Analysis

**DOI:** 10.64898/2026.09.18.26363437

**Authors:** Na Dai, Kirsten L. Waarala

**Affiliations:** College of Osteopathic Medicine, Michigan State University, East Lansing, MI, USA

**Keywords:** Medical education, Osteopathic medical schools, Patient confidentiality, Institutional policies, Decision support, Vignette-based document analysis, Artificial intelligence

## Abstract

**Research Objectives:** To evaluate whether publicly available institutional guidance supports patient-information decisions across scenarios and schools, and characterize document synthesis and evaluation consistency.

**Methods:** We conducted an exploratory, vignette-based document analysis of a geographically diverse nonprobability sample of 20 U.S. osteopathic medical schools. Eight educational vignettes yielded 160 school-vignette pairs. Each pair underwent three separate AI-assisted retrieval-and-evaluation runs, classifying decision support as Explicitly Supported, Inferable, Ambiguous, or Not Addressed. Response selection prioritized greater support for discordant pairs, followed by fewer contributing documents and run order. One investigator verified or revised selected discordant classifications against cited evidence. Explicitly Supported and Inferable were grouped post hoc as sufficient decision support. Analyses were descriptive and included an exploratory two-school model-investigator comparison.

**Results:** At least one eligible source was retrieved for 159 of 160 pairs (99.4%). Final classifications were Explicitly Supported for 18 pairs (11.3%), Inferable for 3 (1.9%), Ambiguous for 132 (82.5%), and Not Addressed for 7 (4.4%). Sufficient support occurred in 21 pairs (13.1%), most frequently for generative AI-assisted reflective writing (7/20 schools, 35%), and in none for personal cloud notes or official clinical logs. Ten schools had no sufficiently supported vignette; the maximum was four of eight. Multiple documents contributed to 111 evaluations (69.4%). Three-run ratings were unanimous for 113 pairs (70.6%), with 80.2% pairwise exact agreement. Investigator review retained 44 of 47 selected discordant ratings and revised three upward. In the two-school comparison, the investigator more frequently judged evidence sufficient when models judged it insufficient than the reverse.

**Conclusions:** Relevant public guidance was frequently retrieved, but few school-vignette pairs were classified as providing sufficient scenario-specific decision support. These findings highlight a gap between identifying relevant guidance and determining an appropriate course of action within this evaluation framework. They support institutional review of how student-facing materials explain information use, storage, sharing, and approval requirements. Vignette-based review identifies questions requiring clarification. Evaluation with learners and assessment of internal and clinical-site guidance would help determine how these findings translate to students’ decisions.

## 1 Background

Educational uses of patient information make confidentiality a practical concern throughout medical training. Students learn through discussing clinical encounters, preparing case presentations, documenting clinical experience, and reviewing clinical images. Research in seven medical schools in England documented students’ use of clinical photographs for learning, while a Canadian study described patient-related communication using students’ personal smartphones [6, 14]. A study of medical students in Spain also documented differences between awareness of confidentiality obligations and reported handling of clinical information [2]. A U.S. survey of medical-school administrators also identified reported incidents of online posting that violated patient confidentiality [4]. These studies connect confidentiality education to concrete information-handling practices, although their different settings and collection periods do not establish current prevalence at U.S. osteopathic schools.

Applying confidentiality responsibilities to an educational activity requires decisions about information content, audiences, systems, and approval. An internal case presentation, a personal study resource, an official clinical log, and an external poster involve different purposes and circumstances. Students may need to determine which details can be included, where materials may be stored, with whom they may be shared, and whether further authorization is needed. Personal digital tools and generative AI introduce additional settings in which patient-derived information might be processed outside established educational systems. A multicenter survey in Ontario documented medical students’ use of generative AI for educational and clinical learning tasks [13]. A French survey of students from several health professions further documented self-reported processing of real medical content during clinical placements, including content students perceived as anonymized [11]. These findings make the handling of patient-derived material in external tools a concrete educational concern. A survey of U.S. osteopathic medical schools documented limited reported development of generative AI policies and training among responding deans and student government presidents [8]. Guidance for educational uses of patient information therefore warrants examination across both longstanding activities and newer technologies.

Previous studies of institutional guidance have examined policy availability and content in domains such as social media and generative AI. An analysis of publicly available policies at U.S. medical schools distinguished general student guidance from explicit instructions about social-media conduct [9]. More recently, a study of AI policies identified reliance on broader university guidance and differences in medical-student specificity [10]. A separate framework-based analysis found uneven coverage of policy topics in AI-related documents across U.S. medical-school institutions [12]. Beyond medical schools, research with university students and staff in Hong Kong informed an AI-policy framework encompassing pedagogical, governance, and operational dimensions, including privacy, infrastructure, and training [3]. These studies provide complementary perspectives on policy availability, content, and implementation, while leaving a distinct question about how guidance resolves specific educational decisions.

Institutional policies may establish privacy requirements without clearly explaining how those requirements apply to specific educational activities. For students using patient information in presentations, assignments, clinical logs, or independent learning, relevant guidance must be interpreted in relation to the proposed activity. Identifying a policy or a statement about confidentiality does not, by itself, establish an appropriate course of action. Conversely, a clear prohibition or an applicable requirement to obtain approval before proceeding may resolve the immediate decision. Document analysis offers a means of studying such institutional materials; the CARDA framework emphasizes transparent reporting of why documents are selected and how they are identified and analyzed in health professions education research [5]. Applying standardized vignettes to retrieved guidance provides a way to examine the distinction between policy availability and decision support.

We evaluated the decision support provided by publicly available policies and training materials at 20 selected U.S. osteopathic medical schools across eight standardized educational vignettes. This exploratory, cross-sectional document analysis used repeated AI-assisted evaluations and targeted investigator review to examine whether retrieved guidance supported an appropriate course of action. The analysis concerned retrieved public materials rather than the completeness of institutional guidance or students’ understanding and behavior.

This work makes three contributions.

1. It evaluates whether publicly available institutional guidance supports an appropriate course of action across eight concrete educational scenarios involving patient information, extending assessment beyond policy availability and topic coverage.
2. It identifies which decision-support elements are resolved and which remain uncertain, distinguishing applicability, permission, information content, storage, sharing, approval, and consultation pathways.
3. It characterizes the consistency and interpretive uncertainty of an AI-assisted evaluation workflow through repeated evaluations, targeted investigator review, and an exploratory comparison between models and investigator ratings.

## 2 Methods

### 2.1 Study Design and Sample

Document analysis provides a systematic approach to examining documents as research data in health professions education [5]. We conducted an exploratory, cross-sectional, vignette-based document analysis to examine whether publicly available institutional guidance supported educational decisions involving patient information. The analysis included policies and training materials from U.S. colleges of osteopathic medicine and focused on whether retrieved guidance supported an appropriate course of action in specified scenarios.

A geographically diverse nonprobability sample of 20 schools was selected with assistance from an OpenAI language model. No formal within-region selection criteria were applied. The sample was intended to provide geographic diversity rather than national representativeness. The included schools, their locations, and regional assignments are listed in Supplementary Table 1.

Each school was evaluated against eight standardized hypothetical vignettes, yielding 160 school– vignette pairs. Document retrieval and evaluation were conducted from August 1 to August 6, 2026. The analysis was restricted to publicly accessible materials and did not assess the completeness of internal institutional guidance, students’ understanding, or their behavior.

### 2.2 Vignettes and Decision-Support Framework

Standardized hypothetical vignettes provided concrete contexts for evaluating whether institutional guidance supported an appropriate course of action. Vignette methodology allows judgments about actions to be examined within specified circumstances [1]. In this study, vignettes were used to evaluate retrieved guidance rather than to elicit students’ attitudes or predict their behavior. The eight activities were an internal case presentation, personal cloud-based learning notes, an official clinical encounter and procedure log, institutional email communication with a supervisor, a small-group clinical discussion, a clinical image screenshot for personal study, an external case-based poster presentation, and generative AI-assisted reflective writing.

The vignettes were developed and refined for clarity, plausibility, and relevance to medical students’ educational use of patient information. N.D. developed the initial drafts with assistance from `gpt-5.6-sol`, after which two authors (N.D. and K.W.) reviewed and iteratively refined them. Each vignette portrayed a student considering an intended activity and described questions that remained unresolved after reviewing institutional guidance. Identical vignette texts were used across schools. Complete texts are provided in Supplementary Material S1.

The evaluation framework comprised eight decision-support elements addressing practical questions raised by the activities. These elements were applicability of guidance to the scenario; whether the activity was permitted; what information could be used; what information must not be used; permitted storage locations; permitted sharing recipients; requirements for additional approval; and pathways for seeking further guidance. The elements informed an overall classification rather than a numerical composite score.

Overall decision support was classified according to whether an appropriate course of action could be identified from the retrieved guidance:

1. **Explicitly Supported**: The appropriate action was readily identifiable with minimal interpretation.
2. **Inferable**: The scenario was not directly addressed, but an appropriate action could reasonably be inferred.
3. **Ambiguous**: Relevant guidance allowed multiple reasonable interpretations.
4. **Not Addressed**: Little or no meaningful guidance relevant to the scenario was identified in the retrieved materials.

For descriptive analysis, Explicitly Supported and Inferable were combined post hoc as “sufficient decision support” during review of the results. This grouping indicated that an appropriate course of action could be determined, regardless of whether the proposed activity was permitted. An applicable prohibition or a clear requirement to obtain approval before proceeding could therefore provide sufficient decision support.

### 2.3 Document Retrieval and Evaluation

AI-assisted searches identified potentially relevant materials on official institutional websites. The evaluation prompt specified that eligible sources must be publicly accessible, relevant to the assigned vignette, and student-facing or expressly applicable to medical students. Eligible source types included student handbooks, catalogs, clinical education manuals, privacy and confidentiality guidance, professionalism policies, technology and artificial intelligence guidance, and official frequently asked questions. University-wide materials and materials from other schools, programs, or clinical affiliates were eligible only when they expressly applied to students at the assigned college or to all university students. Other materials were to be treated as contextual sources or recorded as limitations.

The retrieval instructions required source inspection and restricted evaluations to applicable evidence. Each search query was to include the institution name and relevant terms from a predefined list. The model was instructed to open and inspect sources rather than rely solely on search-result snippets. Inaccessible materials, including those requiring authentication, were to be recorded as limitations and excluded from evidence supporting classifications. The model was instructed not to fill gaps using general privacy knowledge or another institution’s policies. Failure to retrieve guidance was not interpreted as evidence that no internal policy existed, and guidance allowing multiple reasonable interpretations was to be classified as Ambiguous.

Each school–vignette pair underwent three separate evaluations using `gpt-5.6-luna` through the OpenAI Responses API with the `web_search` tool enabled. Requests were submitted sequentially using identical instructions and vignette text, without passing previous responses into subsequent requests. Reasoning effort, temperature, seed, and output-token limits were not explicitly configured. Each evaluation returned a structured response containing an overall classification, rationale, decision-support answers, source references, document counts, and limitations. The full prompt and output schema are provided in Supplementary Material S2.

Document counts distinguished inspected sources from sources that materially contributed to the evaluation. The prompt defined a document as one unique official webpage or downloadable document; separate pages within the same PDF, duplicate URLs, search-result pages, and repeated access to the same source were not to be counted separately. A document was considered materially contributing if it changed the overall classification or directly answered at least one decision-support element. Counts were reported by the model in its structured output.

One response was selected for each school–vignette pair using predefined selection rules. For pairs with unanimous classifications, selection prioritized the fewest contributing documents, followed by run order to resolve ties. For discordant pairs, selection prioritized classifications in the order Explicitly Supported, Inferable, Ambiguous, and Not Addressed, followed by the fewest contributing documents and run order. Thus, selection for discordant pairs prioritized the classification indicating greater decision support rather than the majority classification.

Targeted investigator review assessed the selected response for each pair with discordant model classifications. N.D. opened the cited source documents and inspected the extracted passages to determine whether the selected classification was supported. Unsupported classifications were directly revised using the decision-support framework. Pairs with unanimous classifications did not undergo this targeted review. The final analytical classification incorporated investigator corrections where applicable.

### 2.4 Outcomes and Analysis

The primary analytical unit was the school–vignette pair, with 160 pairs clustered within 20 schools. The primary outcome was the final four-category decision-support classification. Counts and percentages were summarized overall and by vignette, together with the post hoc sufficient-support grouping. School-level summaries described the number of vignettes with sufficient decision support out of eight evaluated scenarios.

Source coverage and document counts characterized the retrieval-and-evaluation workflow. We summarized whether at least one eligible source was reported across the three evaluations for each pair. Document counts from selected responses were summarized using medians and interquartile ranges, together with the frequencies of zero, one, or multiple contributing documents. These measures described the workflow and were not interpreted as measures of student effort.

Exploratory element-level coding examined which practical questions were resolved within the selected responses. Applicability retained its original yes, no, or unclear classification. Answers to the remaining seven elements were assigned one of six statuses: resolved, partially resolved, unresolved, conflicting, not applicable, or uncodable. Coding considered each answer in relation to the vignette and companion answers rather than relying solely on keywords or the overall classification. Clear permission, prohibition, or an applicable approval requirement could resolve an element. Status frequencies were summarized separately for each element, retaining all 160 pairs in each denominator and reporting not-applicable and uncodable answers separately. Previous research has examined the use of large language models with explicit codebooks to support qualitative coding [15]. Definitions and boundary rules are provided in Supplementary Material S3.

Qualitative content analysis involves interpreting textual material through categories that organize its meaning [7]. We used exploratory content coding to characterize uncertainty expressed in the model-reported limitations of selected evaluations. Limitation statements were considered together within each school–vignette response and coded using a framework organized around access and availability, applicability and currency, operational clarity, and scenario-specific interpretation and synthesis. Multiple themes could be assigned to a response, but each theme was counted at most once per school–vignette pair. Frequencies therefore represented the number of pairs reporting a theme rather than the number of individual statements. N.D. approved the element-level and reported-limitations coding frameworks. Individual code assignments were AI-assisted and retained without item-by-item investigator verification. The reported-limitations codes characterized uncertainty expressed in the evaluations and did not independently establish institutional policy deficiencies. The reported-limitations framework is provided in Supplementary Material S4.

Consistency across repeated evaluations was assessed using original model classifications before investigator corrections. We calculated the proportion of pairs with unanimous three-run classifications and pairwise exact agreement. Each pair contributed three comparisons between runs, yielding 480 comparisons across 160 pairs. Pairwise exact agreement was the number of matching comparisons divided by the total number of comparisons. Repeated evaluations assessed workflow consistency, and comparisons within pairs were not treated as independent observations.

Targeted investigator-review findings were summarized separately from model consistency. We reported the number of selected classifications retained or revised and the direction of revisions. Because review was restricted to pairs with discordant model classifications, these findings were not interpreted as estimates of accuracy across the full sample.

An exploratory comparison examined two randomly selected schools, AZCOM and MSUCOM, using both `gpt-5.6-luna` and `gpt-5.6-sol`. Both models used identical retrieval instructions, vignette texts, tools, and execution settings. Each model contributed three responses for each of eight vignettes per school, yielding 16 school–vignette pairs and 48 responses per model. Comparisons used original model classifications. Pair-level agreement was calculated using each model’s majority classification across its three runs. For sufficient/insufficient comparisons, individual run classifications were grouped before determining the binary majority. Agreement across all nine cross-model run combinations within each pair was also summarized.

Response-specific investigator ratings provided an additional exploratory comparison with model classifications. N.D. interpreted the evidence supplied in each model-generated response and assigned a decision-support classification, with instructions to base judgments on that evidence without supplementing it with outside knowledge. Each investigator rating was compared with the corresponding original model classification. Unrecorded investigator ratings were excluded from the relevant comparisons and reported. Because the ratings concerned model-specific evidence bundles, they were not treated as a common independent reference standard.

Agreement analyses included exact four-category agreement, sufficient/insufficient agreement, and a post hoc within-one-category sensitivity analysis. The latter counted identical or adjacent classifications in the order Explicitly Supported, Inferable, Ambiguous, and Not Addressed. This ordering was an analytical convention and did not imply that category distances were empirically calibrated or that adjacent classifications were interchangeable. All analyses were descriptive, and repeated responses and comparisons within school–vignette pairs were not treated as independent observations.

Analyses were performed in Python 3 within Google Colab. OpenAI Codex assisted with writing and revising the analysis code.

### 2.5 Ethics

The study analyzed publicly accessible institutional materials using hypothetical educational scenarios. No participants were recruited, and no identifiable private information was collected or analyzed. The authors considered the study not to constitute human subjects research; therefore, institutional review board review was not sought.

## 3 Results

### 3.1 Study Coverage and Overall Decision Support

At least one eligible source was identified across the three evaluations for 159 of 160 school– vignette pairs (99.4%). In the final classification dataset, 18 pairs (11.3%) were classified as Explicitly Supported, 3 (1.9%) as Inferable, 132 (82.5%) as Ambiguous, and 7 (4.4%) as Not Addressed. Under the post hoc grouping of Explicitly Supported and Inferable, 21 pairs (13.1%) provided sufficient decision support.

### 3.2 Decision Support Across Vignettes and Schools

Sufficient decision support varied across the eight vignettes (Figure 1). It was most frequent for generative AI-assisted reflective writing (7/20 schools, 35.0%), followed by institutional email communication with a supervisor and external case-based poster presentation (4/20 each, 20.0%). Internal case presentation, small-group clinical discussion, and clinical image screenshots for personal study each had sufficient support at 2/20 schools (10.0%). No school provided sufficient support for personal cloud-based learning notes or the official clinical log. Ambiguous was the most frequent classification for every vignette, ranging from 13/20 schools (65.0%) for generative AI-assisted reflective writing to 20/20 (100.0%) for the official clinical log.

**Figure 1:**
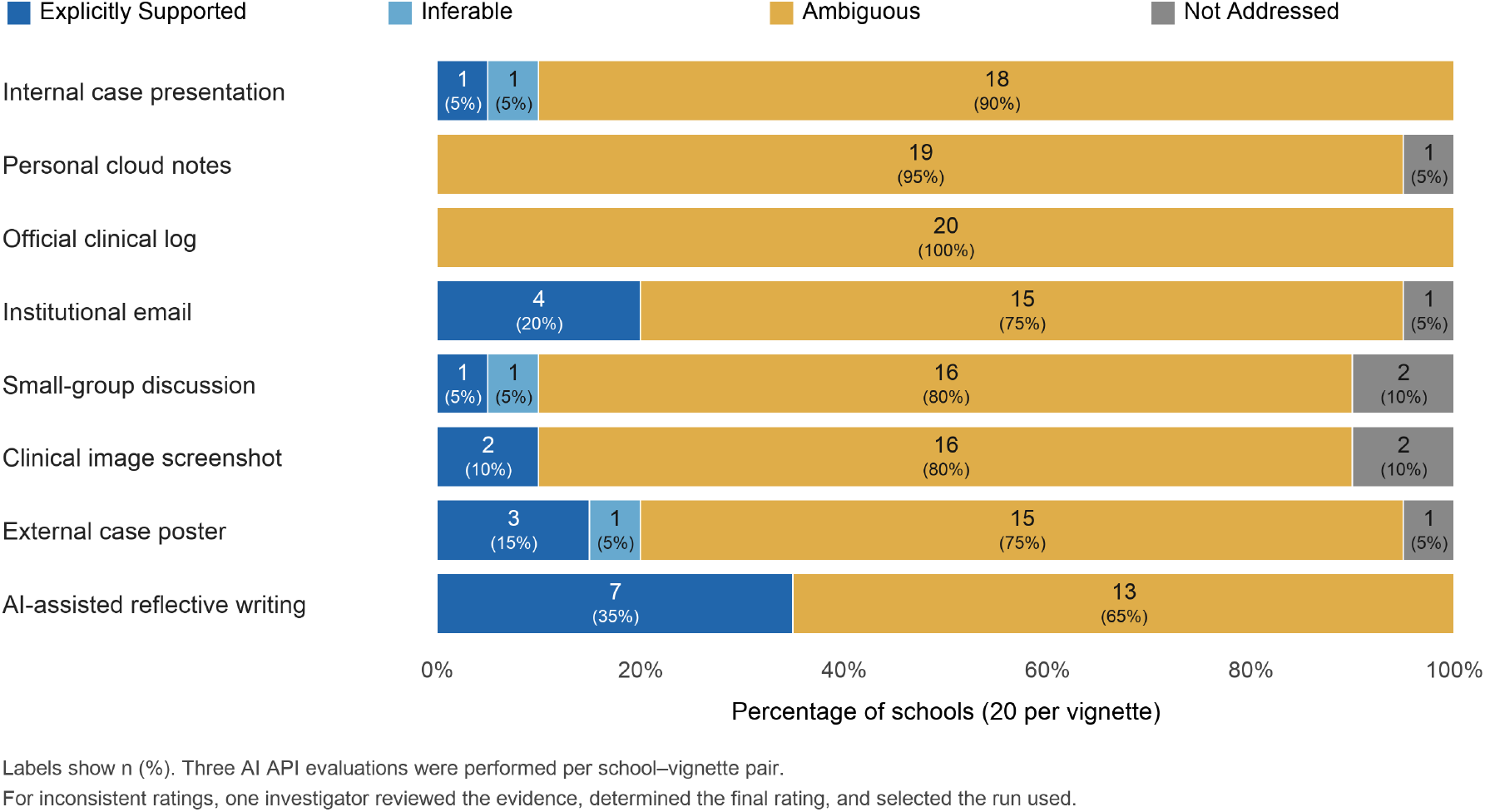
Final decision-support classifications across eight educational vignettes. Each bar represents 20 schools; segment labels show counts and percentages. Each school–vignette pair underwent three AI-assisted evaluations. One response per pair was selected using the specified classification-priority and tie-breaking rules. For pairs with discordant classifications, one investigator reviewed the selected response’s cited evidence and retained or revised its classification.

Across schools, 10 of 20 (50.0%) provided sufficient decision support for at least one vignette. The number of sufficiently supported vignettes ranged from zero to four per school. School-level classifications across all eight vignettes are presented in Supplementary Figure 1.

### 3.3 Decision Support Across Individual Elements

All 160 selected school–vignette responses contained answers to the eight decision-support questions. Question 1 assessed applicability and was summarized separately using its original categories: yes in 95 responses (59.4%), no in 12 (7.5%), and unclear in 53 (33.1%). Questions 2–8 assessed distinct aspects of decision resolution.

In the current AI-assisted contextual coding, resolved decision support was most frequent for identifying where to seek further guidance (Q8; 95/160, 59.4%). The corresponding counts were 33/160 (20.6%) for identifying permitted sharing recipients (Q6), 21/160 (13.1%) for determining permitted storage or processing locations (Q5), 19/160 (11.9%) for determining whether the activity was permitted (Q2), 12/160 (7.5%) for identifying information that should not be used (Q4), 11/160 (6.9%) for identifying information that may be used (Q3), and 8/160 (5.0%) for determining whether additional approval was required (Q7). Full classification distributions are presented in Table 1, and vignette-specific resolved counts are shown in Figure 2.

**Table 1:** Decision-resolution classifications for Questions 2–8 under AI-assisted contextual coding.

| Question | Resolved | Partially resolved | Unresolved | Conflicting | Not applicable | Uncodable |
| --- | --- | --- | --- | --- | --- | --- |
| Q2: Activity permission | 19 (11.9) | 19 (11.9) | 122 (76.2) | 0 (0.0) | 0 (0.0) | 0 (0.0) |
| Q3: Permissible information | 11 (6.9) | 19 (11.9) | 130 (81.2) | 0 (0.0) | 0 (0.0) | 0 (0.0) |
| Q4: Excluded information | 12 (7.5) | 22 (13.8) | 124 (77.5) | 1 (0.6) | 0 (0.0) | 1 (0.6) |
| Q5: Storage or processing | 21 (13.1) | 0 (0.0) | 119 (74.4) | 0 (0.0) | 20 (12.5) | 0 (0.0) |
| Q6: Sharing recipients | 33 (20.6) | 10 (6.2) | 97 (60.6) | 0 (0.0) | 20 (12.5) | 0 (0.0) |
| Q7: Additional approval | 8 (5.0) | 12 (7.5) | 140 (87.5) | 0 (0.0) | 0 (0.0) | 0 (0.0) |
| Q8: Further guidance | 95 (59.4) | 32 (20.0) | 33 (20.6) | 0 (0.0) | 0 (0.0) | 0 (0.0) |

Values are n (%) of 160 school-vignette responses per question. Question numbers denote decision-support elements, not vignette numbers. All six statuses are retained, including not-applicable and uncodable answers in the denominator. Partially resolved is not combined with resolved. Q1 uses its original yes/no/unclear scale and is reported separately.

**Figure 2:**
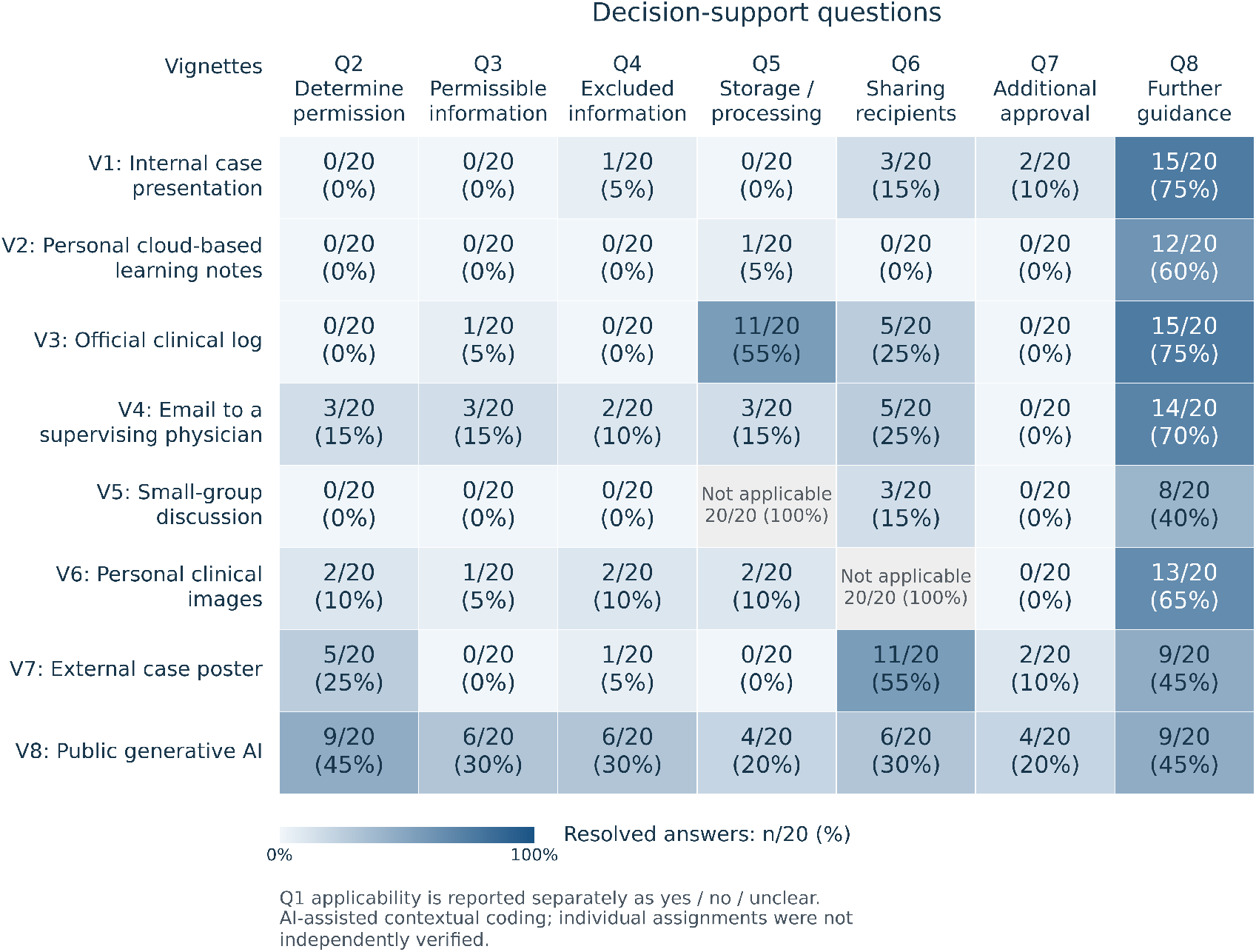
Decision resolution by vignette and question. Rows are the eight educational vignettes (V1–V8); columns are the seven decision-resolution questions (Q2–Q8). Blue cells show resolved answers as n/20 (%) across schools. Gray cells denote a question that does not arise in the specified vignette, rather than absent guidance. Q1 applicability is summarized separately. Counts reflect AI-assisted contextual coding retained without item-by-item investigator verification.

For Q2, resolution indicated that an appropriate course of action could be determined, including a prohibition or an explicit requirement to await approval; it did not necessarily indicate permission to undertake the proposed activity.

### 3.4 Reported Limitations of Document Retrieval and Evaluation

All 160 selected school–vignette responses contained reported limitations, comprising 730 statements. These statements were coded through AI-assisted contextual analysis within their respective school–vignette passages using a coding framework approved by N.D. (Supplementary Material S4). Of the 730 statements, 711 contained substantive limitation content and 19 contained only procedural or contextual information. Each detailed theme was counted once per school–vignette pair, irrespective of repeated mentions.

Reported limitations were organized into four supercategories: access and availability of guidance; applicability and currency of guidance; operational clarity of guidance; and scenario and synthesis constraints (Figure 3).

**Figure 3:**
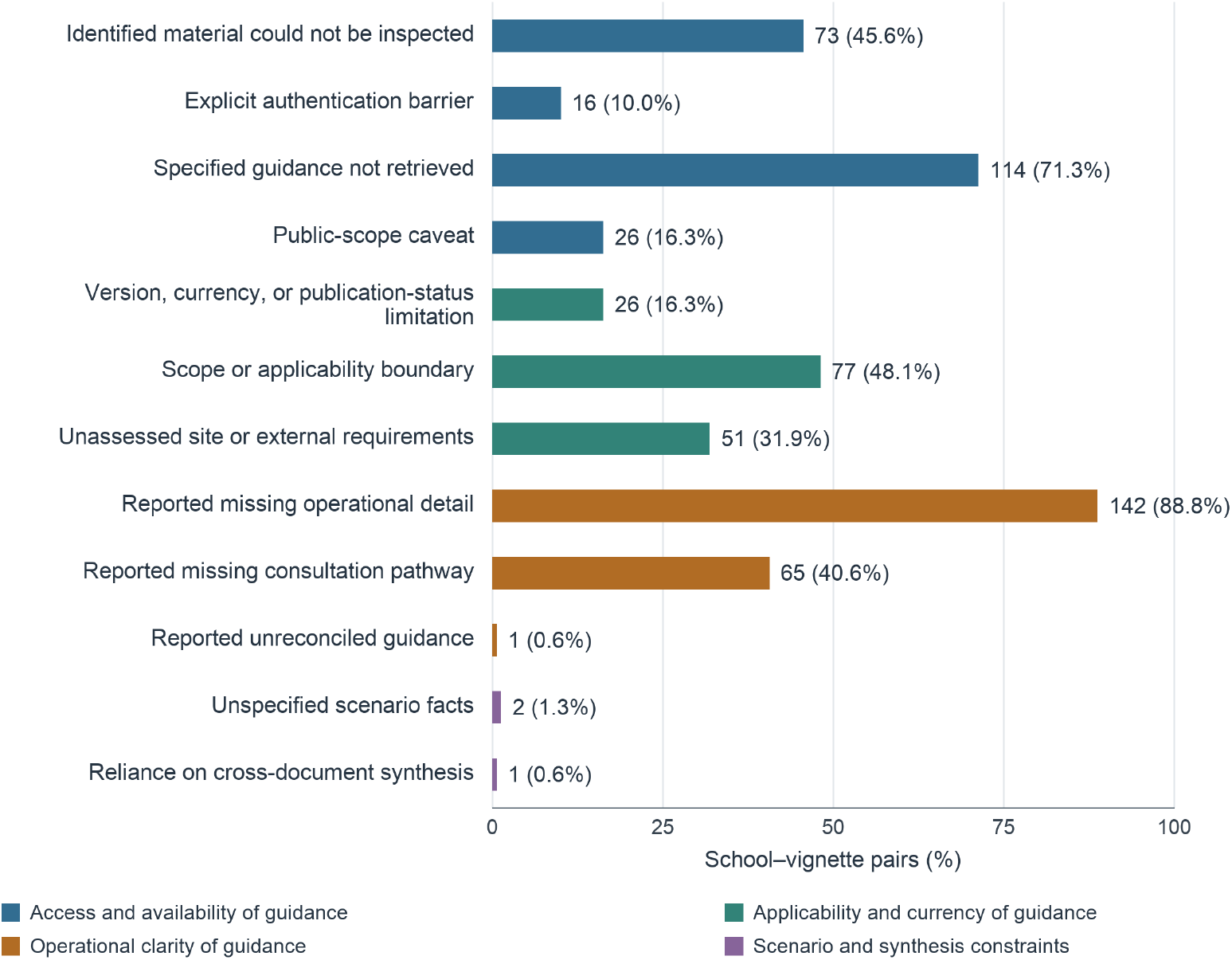
Reported limitations across 160 selected school–vignette evaluations. Bars represent detailed codes and show counts and percentages using 160 as the denominator. Colors identify four supercategories: access and availability of guidance (blue), applicability and currency of guidance (teal), operational clarity of guidance (orange), and scenario and synthesis constraints (purple). Each theme was counted once per pair; themes could overlap. Procedural or contextual-only statements were excluded from theme counts. Findings describe reported evaluation limitations rather than independently verified policy deficiencies. Coding was AI-assisted using an investigator-approved framework.

Within access and availability, specified guidance was not retrieved for 114 pairs (71.3%), and identified materials could not be inspected for 73 (45.6%). Explicit authentication barriers were reported for 16 pairs (10.0%), while 26 (16.3%) included caveats about potentially available internal or otherwise unobserved guidance. Regarding applicability and currency, scope or applicability boundaries were reported for 77 pairs (48.1%), unassessed site or external requirements for 51 (31.9%), and version, currency, or publication-status concerns for 26 (16.3%).

Missing operational detail was the most frequently coded theme, occurring in 142 pairs (88.8%). Missing consultation pathways were reported for 65 pairs (40.6%), and unreconciled guidance for one (0.6%). Unspecified scenario facts and reliance on cross-document synthesis were reported for two pairs (1.3%) and one pair (0.6%), respectively. The cross-document synthesis theme captured explicit statements about reliance on multiple sources within the limitations narratives; it was not assigned solely because multiple documents contributed to an evaluation. These themes characterize the limitations reported by the selected evaluations; an identified omission did not necessarily prevent determination of an appropriate immediate action.

### 3.5 Document Review and Source Contributions

Across the 160 selected school–vignette evaluations, the median number of documents reported as reviewed was 4 (interquartile range [IQR], 4–6), while the median number reported as materially contributing was 2 (IQR, 1–3). Material contribution referred to documents that informed the overall classification or directly answered at least one decision-support question.

Multiple materially contributing documents were reported for 111 pairs (69.4%), and a single contributing document for 44 pairs (27.5%). This document-count measure captures the use of multiple sources regardless of whether cross-document synthesis was explicitly described in the limitations narrative. The remaining five selected evaluations (3.1%) reported no materially contributing document. These counts describe the selected evaluation for each pair, whereas the source-coverage summary in Section 3.1 considered all three evaluations. Document counts characterize evidence use within the evaluation workflow rather than the effort required of students.

### 3.6 Consistency Across Evaluations and Human Review

Repeated Luna evaluations produced unanimous four-category classifications for 113 of 160 school–vignette pairs (70.6%); 47 pairs (29.4%) were discordant. Pairwise exact agreement was 385/480 within-pair run comparisons (80.2%).

Targeted investigator review retained the selected classification for 44 of the 47 discordant pairs (93.6%) and revised three (6.4%): one from Ambiguous to Explicitly Supported and two from Ambiguous to Inferable. These findings describe review of selected responses from discordant pairs and do not estimate accuracy across the full sample.

In the exploratory AZCOM–MSUCOM subset, comprising 16 pairs and 48 responses per model, Luna and Sol agreed on majority four-category classifications for 12/16 pairs (75.0%) and sufficient/insufficient classifications for 14/16 (87.5%; Table 2). Both models classified one pair as sufficient and 13 as insufficient. For the two binary disagreements—AZCOM vignette 6 and MSUCOM vignette 8—Luna classified support as sufficient and Sol as insufficient.

**Table 2:** Agreement between Luna and Sol in the exploratory two-school subset.

| Agreement criterion | Majority ratings | All run combinations |
| --- | --- | --- |
| Exact four-category | 12/16 (75.0) | 98/144 (68.1) |
| Sufficient versus insufficient | 14/16 (87.5) | 123/144 (85.4) |
| Within one category | 14/16 (87.5) | 127/144 (88.2) |

Values are n/N (%), using original model classifications. The AZCOM-MSUCOM subset included 16 school-vignette pairs, with three runs per model per pair. All-run comparisons include nine cross-model combinations per pair. Sufficient support combines Explicitly Supported and Inferable; binary grouping preceded majority determination. Within-one-category agreement is a post hoc measure allowing identical or adjacent classifications in the order Explicitly Supported, Inferable, Ambiguous, and Not Addressed. Repeated comparisons are dependent.

Exact and binary agreement percentages with the investigator’s response-specific ratings were lower than the corresponding model–model percentages. Investigator ratings were available for 47 Luna responses and all 48 Sol responses. Exact agreement was 20/47 (42.6%) for Luna and 18/48 (37.5%) for Sol; binary agreement was 30/47 (63.8%) and 26/48 (54.2%), respectively (Table 3). The investigator rated evidence sufficient when the model rated it insufficient for 15 Luna and 17 Sol responses; the reverse occurred for two and five responses, respectively. These ratings were separate from the targeted review of discordant pairs and concerned potentially different model-supplied evidence bundles, rather than a common independent reference standard.

**Table 3:**
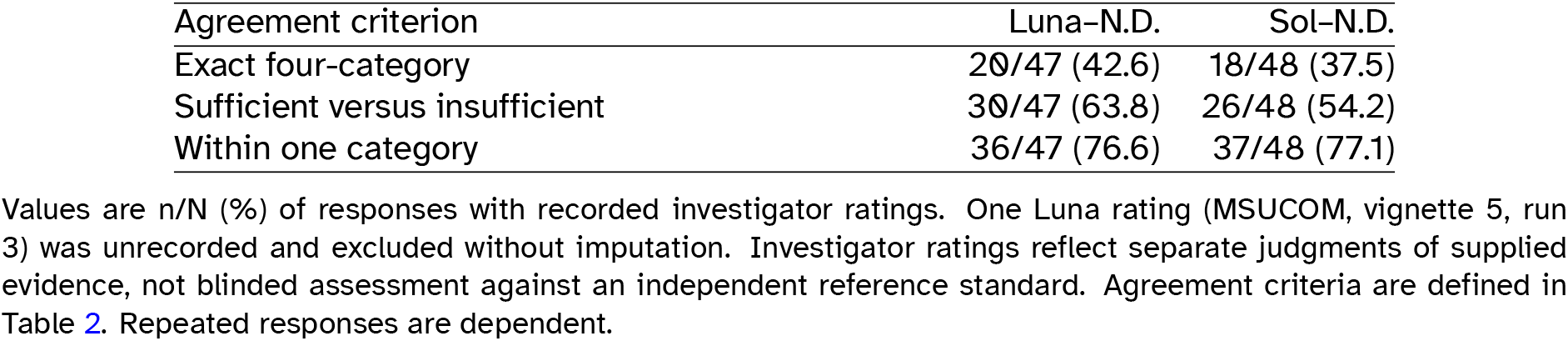
Agreement between model classifications and N.D.’s response-specific ratings of supplied evidence.

| Agreement criterion | Luna–N.D. | Sol–N.D. |
| --- | --- | --- |
| Exact four-category | 20/47 (42.6) | 18/48 (37.5) |
| Sufficient versus insufficient | 30/47 (63.8) | 26/48 (54.2) |
| Within one category | 36/47 (76.6) | 37/48 (77.1) |

Values are n/N (%) of responses with recorded investigator ratings. One Luna rating (MSUCOM, vignette 5, run 3) was unrecorded and excluded without imputation. Investigator ratings reflect separate judgments of supplied evidence, not blinded assessment against an independent reference standard. Agreement criteria are defined in Table 2. Repeated responses are dependent.

Within-one-category agreement was higher than exact agreement in all comparisons (Tables 2 and 3); each model differed from the investigator by at least two categories for 11 responses. However, this sensitivity measure permits agreement between Inferable and Ambiguous, which lie on opposite sides of the sufficient-support threshold. It therefore does not establish agreement about whether guidance provided sufficient decision support.

## 4 Discussion

### 4.1 Principal findings

In this exploratory analysis of 20 U.S. osteopathic medical schools, at least one eligible public source was retrieved across three evaluations for 159 of 160 school–vignette pairs, but only 21 pairs (13.1%) were ultimately classified as providing sufficient decision support. Most pairs were classified as Ambiguous. This pattern distinguishes finding relevant guidance from determining an appropriate action for a specific educational scenario. Guidance may apply to medical students and address confidentiality without resolving the particular questions about information use, storage, sharing, or approval posed by an activity. Conversely, a clear prohibition or an applicable requirement to obtain approval before proceeding can resolve a decision without permitting the proposed activity.

The selected evaluations frequently reported missing operational detail, uncertainty about applicability, and difficulties accessing identified materials. These reports help characterize the uncertainty encountered when applying retrieved guidance to concrete educational decisions.

The model–investigator comparison further indicates that judgments of decision support depend on how retrieved evidence is interpreted. Evaluation consistency and interpretive differences are therefore integral to understanding the predominance of insufficient-support classifications and the questions identified for institutional review.

### 4.2 Interpretation across scenarios and decision-support elements

Previous studies provide context for the distinction between policy availability and scenario-specific decision support. Ichikawa et al. documented limited reported development of generative AI policies and training among responding deans and student government presidents at U.S. osteopathic medical schools [8]. Knopf et al. identified substantial reliance on broader university AI policies, which were generally less specific to medical students than policies originating from medical schools [10]. Rush et al. further examined the breadth of AI-related guidance across U.S. medical-school institutions, finding uneven coverage across a framework of policy topics [12]. Their assessment of topic coverage differs from our assessment of whether guidance resolves a particular decision. Differences in samples, collection periods, and outcomes preclude direct comparison of their estimates with ours. Our analysis complements this work by examining whether retrieved guidance resolves defined educational decisions involving patient information, including activities beyond AI use.

We conjecture that the greater frequency of sufficient classifications for generative AI-assisted reflective writing, compared with none for personal cloud notes or official clinical logs, may partly reflect differences in when guidance developed and the channels through which it is disseminated, alongside differences in content. Guidance for a newly emerging activity such as generative AI use may be developed and disseminated through publicly accessible university or technology-office webpages. Instructions for longstanding activities may remain embedded in established channels, including clerkship orientation, internal manuals, supervision, and restricted learning-management systems. When and how guidance developed could therefore influence its visibility to a public-web search. This explanation remains tentative because the study did not systematically examine dissemination channels or internal resources. The AI findings also should not be read as evidence of greater permission to use these tools, because clear restrictions can provide sufficient decision support.

Support also varied across schools: half had at least one vignette classified as sufficiently supported, and school-level counts ranged from zero to four of the eight vignettes. This variation supports examining guidance in relation to a particular activity rather than treating decision support as a uniform institutional characteristic. Differences may arise from the materials retrieved, their applicability to the scenario, and their interpretation within the workflow.

The AI-assisted element-level coding illustrates how selected responses resolved different parts of a scenario independently. In the selected PNWU-COM clinical-log response, the designated clinical logging system (eValue) resolved the storage question (Q5), while the permissibility of all proposed fields, including date of service, remained unresolved (Q3). In the selected BCOM supervisor-email response, participating providers were identified as recipients for direct patient-management communication, resolving the recipient question (Q6), but the acceptability of the proposed combination of initials, age range, inpatient unit, and laboratory result remained unresolved (Q3). In the selected ACOM external-poster response, the Office of Research provided an identified route for compliance and procedural assistance (Q8), while the applicable approval requirements remained unresolved (Q7); several relevant linked resources were reported as inaccessible.

Together, these examples distinguish knowing where to store information, with whom to share it, and whom to consult from knowing which information is permissible or which approvals are required. The relatively frequent resolution of consultation pathways suggests that retrieved guidance can identify a useful next step without resolving the substantive decision. Under the framework, an explicit, applicable instruction to obtain approval before proceeding can resolve the immediate action: the student must await approval. Identifying a general contact alone may leave that action unresolved. Resolution of the consultation element therefore does not automatically establish sufficient support for the whole scenario.

### 4.3 Evaluation consistency and investigator disagreements

Repeated evaluations showed consistency, although its extent depended on the measure used. All three original ratings agreed for 113 of 160 pairs (70.6%), whereas 385 of 480 comparisons between two runs agreed exactly (80.2%). The latter measure also counts matching runs within otherwise discordant pairs. These findings describe repeatability within the workflow, not the correctness of the classifications: shared retrieval constraints or interpretive tendencies may persist across runs. Targeted investigator review retained 44 of 47 selected ratings from discordant pairs, but this retention rate cannot validate the full sample. The implications of this targeted review and the response-selection procedure are considered in Section 4.5.

The separate two-school comparison showed that agreement between models can coexist with substantive disagreement with an investigator. Binary majority classifications agreed between models for 14 of 16 pairs, including 13 pairs classified as insufficient by both. Agreement with N.D.’s response-specific interpretation of the supplied evidence was lower. The investigator judged evidence sufficient when the model judged it insufficient for 15 Luna and 17 Sol responses, whereas the reverse occurred for two and five responses, respectively. This direction of disagreement warrants attention when interpreting the predominance of insufficient-support classifications, but it does not establish model error or a correction that can be applied to the full sample.

We also conjecture that prior knowledge and familiarity with clinical education may have implicitly influenced the investigator’s interpretation, despite instructions to base ratings on the supplied evidence without using outside knowledge to fill gaps. Such knowledge could connect general guidance to a proposed activity, making an appropriate action appear reasonably inferable despite limited explicit documentary support. This offers one possible explanation for the investigator’s greater tendency to assign sufficient-support ratings. However, the study did not directly assess the use of background knowledge by either the investigator or the models and cannot attribute the disagreements to this mechanism or assume that models excluded such knowledge more successfully. The findings highlight the need to clarify the boundary between an inference supported by the supplied evidence and an assumption introduced by the evaluator.

Higher within-one-category agreement does not remove the substantive disagreement about sufficient support. Inferable and Ambiguous are adjacent categories but fall on opposite sides of that threshold. Exact, binary, and adjacent-category agreement therefore address different questions. Taken together, the comparisons indicate that consistency should be considered alongside differences in interpretation when assessing the decision support attributed to retrieved guidance.

### 4.4 Practical implications

The limitations reported in Figure 3 identify priorities for further institutional review. Missing operational detail was the most frequent theme, accompanied by difficulties retrieving specified guidance, determining applicability, inspecting identified materials, and locating consultation pathways. These reported difficulties suggest concrete questions for examining how guidance is organized, communicated, and applied to educational activities.

First, institutions could examine whether guidance resolves the operational decisions involved in common educational activities. Scenario-based examples could connect general confidentiality requirements to permissible information, approved storage or processing systems, permitted recipients, and approval requirements. Examples should make the appropriate immediate action clear, including when an activity must be deferred or avoided. The purpose would be to answer the questions necessary for the activity rather than provide exhaustive detail about every possible circumstance.

Second, institutions could review how students locate relevant guidance and determine its applicability. A central index or clear links between public webpages, handbooks, clerkship materials, and internal learning systems could help students identify the relevant resources. Materials could specify the students, activities, and settings they cover and explain how school guidance relates to clinical-site requirements. Accessibility should be assessed through the accounts and systems available to enrolled students, because a failed public-web retrieval does not establish that students lack access. Improving discoverability may therefore involve connecting existing resources rather than making all guidance public or creating additional policies.

Third, guidance could provide an actionable consultation pathway when written instructions do not resolve a decision. A historical analysis of publicly available social-media policies at U.S. medical schools described both explicit behavioral restrictions and contacts for students uncertain about appropriate conduct [9]. These features provide precedents for policy design. Identifying an appropriate office or role, the questions it can address, and how to contact it could help students seek assistance. Materials should distinguish a request for advice from a required approval process. When approval is required, guidance should identify the responsible role or office and state that the activity must await approval. When only a consultation contact is provided, guidance should clarify what the student may do while awaiting advice. These distinctions would help students identify an appropriate immediate action without treating a contact listing as authorization.

Finally, these proposed improvements require evaluation with learners. Previous research has documented differences between medical students’ awareness of confidentiality obligations and their reported handling of clinical information, reinforcing the need to assess understanding and practice separately [2]. A survey at a Canadian medical school also documented patient-related smartphone communication alongside concerns about confidentiality [14]. These findings support examining how students locate, interpret, and apply guidance to communication decisions. Students at different training stages could be asked to locate relevant guidance, explain its applicability, and identify an appropriate action for standardized scenarios. Observing their searches and reasoning could help distinguish difficulties with access, unclear instructions, and assumptions informed by prior experience. Comparing these assessments before and after revisions would help determine whether changes improve decision support.

### 4.5 Strengths and limitations

Strengths of this study include the application of standardized educational vignettes across schools, repeated evaluations, and explicit frameworks for overall decision support, individual elements, and reported evaluation limitations. Preserved source references, original responses, and coding decisions support traceability of the analysis. Targeted investigator review examined the cited evidence for discordant pairs, while the separate model–investigator comparison revealed interpretive differences not captured by repeated model evaluations alone. Together, these features provide a structured approach to exploring how retrieved institutional guidance addresses concrete educational decisions.

The sample and scenarios limit the scope of inference. School selection was nonprobability and model-assisted, with geographic diversity rather than national representativeness as its objective. The findings therefore do not support national prevalence estimates or comparisons of regional policy quality. The eight vignettes represent a limited set of educational activities and information-handling decisions. Their explicit descriptions of unresolved questions may have influenced model interpretation, and different scenario wording or details could yield different classifications.

The public-source restriction and AI-assisted workflow also constrain the findings. Internal materials, orientation, supervision, informal consultation, and clinical-site guidance were not comprehensively assessed. Failure to retrieve or open a source may reflect search coverage or tool-access constraints rather than its absence or inaccessibility to enrolled students. Retrieval and interpretation depended on the model and web-search tools available through the API during the study period. Repeated evaluations could share omissions or interpretive tendencies; agreement across runs therefore does not establish the completeness of retrieval or correctness of the resulting classifications.

Review and analytical procedures impose further limitations. Targeted source review covered discordant pairs, while unanimous pairs did not receive that review. Prioritizing the highest-support response for discordant pairs may favor sufficient-support classifications relative to majority-based selection; the final results should therefore be interpreted in relation to that procedure. Element and limitation analyses coded model-generated content. N.D. approved both coding frameworks, but individual element and reported-limitations code assignments were not independently verified by an investigator and may contain classification errors. Framework approval also did not constitute verification of the underlying sources. The coded themes describe limitations reported in the evaluations rather than independently established policy deficiencies; a reported omission did not necessarily prevent determination of an appropriate immediate action. The sufficient-support grouping and within-one-category comparison were post hoc. Observations were clustered within schools and school–vignette pairs, and repeated comparisons were dependent. The exploratory comparison included only two schools and one investigator, who interpreted the potentially different evidence supplied in each output rather than independently retrieving a common reference set for each pair. One investigator rating was unrecorded and excluded without imputation. These comparisons cannot establish model accuracy against an independent reference standard or the superiority of one model over another.

Finally, the study did not assess student understanding, behavior, institutional compliance, or patient outcomes. Document counts describe evidence use within the selected evaluations rather than the effort students expend in locating or interpreting guidance. The findings identify questions for further source verification and learner-based evaluation; they do not establish overall institutional policy quality or demonstrate that the proposed changes to guidance would improve educational decisions.

## 5 Conclusions

In this exploratory analysis of 20 U.S. osteopathic medical schools, relevant public guidance was retrieved for nearly all school–vignette pairs, but only 21 of 160 pairs (13.1%) were classified as providing sufficient scenario-specific decision support. These findings highlight a gap between identifying relevant guidance and determining an appropriate course of action within this evaluation framework. They support institutional review of how student-facing materials explain information use, storage, sharing, approval requirements, and consultation pathways. Vignette-based document analysis offers a practical approach to identifying questions requiring clarification and informing the development of scenario-specific guidance. Further source verification, assessment of internal and clinical-site materials, and evaluation with learners would help establish whether the uncertainties identified through AI-assisted review arise in practice and whether revised guidance improves students’ decision-making.

## Data Availability

All institutional source materials analyzed in this study were publicly accessible at the time of data collection. The derived data supporting the findings of this study, including the AI-assisted evaluation outputs, classifications, and coding data, are available from the corresponding author upon reasonable request. The evaluation prompt, coding frameworks, and study vignettes are provided in the Supplementary Information.

## Supplementary Information

### Supplementary Material S1: Educational Vignettes

The following eight hypothetical vignettes were used to evaluate decision support in publicly available institutional policies and training materials. Identical vignette texts were used across the 20 schools. The wording below reproduces the scenarios used during the study evaluations.

#### Vignette 1: Internal Case Presentation

A conscientious student has reviewed the institution’s guidance.

A third-year medical student is assigned to present an inpatient case during an internal noon conference attended by faculty, residents, and medical students. To explain the diagnosis and clinical reasoning, the student prepares slides containing the patient’s age range, sex, relevant medical history, laboratory and imaging findings, hospital course, and a clinical photograph of a skin lesion. The patient’s name, medical-record number, full date of birth, and face are not included.

Before the presentation, the student reviews the institution’s publicly available privacy and professionalism materials. The student remains uncertain whether the de-identified clinical photograph and accompanying case details may be used in an internal educational presentation, or whether additional approval is required.

#### Vignette 2: Personal Cloud-Based Learning Notes

A conscientious student has reviewed the institution’s guidance.

During a clerkship, a third-year medical student wants to keep a personal set of learning notes to review common diagnoses and teaching points encountered on the wards. The student plans to use a personal cloud-based note application and record each patient’s age range, diagnosis category, key clinical finding, and a brief learning point. The notes will not contain patient names, medical-record numbers, full dates of service, photographs, or other direct identifiers.

Before creating the notes, the student reviews the institution’s publicly available privacy and technology materials. The student remains uncertain whether patient-derived clinical information may be retained in a personal cloud-based tool for educational purposes, whether an institution-approved system is required, and which details must be omitted.

#### Vignette 3: Official Clinical Encounter and Procedure Log

A conscientious student has reviewed the institution’s guidance.

A third-year medical student must enter required clinical encounters and procedures into the institution’s official clerkship logging system. The system asks for the clinical setting, diagnosis category, procedure, supervising clinician, and date of service. To complete the requirement accurately, the student plans to enter information directly into the official system after each encounter and does not intend to retain a separate copy.

Before doing so, the student reviews the institution’s publicly available clerkship, privacy, and technology materials. The student remains uncertain whether the required entry of patient-related clinical information into the official educational logging system is permitted as described, whether any fields should be omitted or generalized, and whom to contact if the system requests more information than seems necessary.

#### Vignette 4: Institutional Email Communication With a Supervisor

A conscientious student has reviewed the institution’s guidance.

During a clerkship, a third-year medical student has a question about a patient’s diagnostic workup and wants to ask the supervising physician for feedback. The student plans to send a message from the student’s institution-issued email account that includes the patient’s initials, age range, inpatient unit, relevant laboratory result, and a brief description of the clinical question. The message would be sent only to the supervising physician.

Before sending the message, the student reviews the institution’s publicly available privacy, communication, and technology materials. The student remains uncertain whether this information may be sent by institutional email, whether a more secure clinical messaging system is required, and whether the patient details should be further limited.

#### Vignette 5: Small-Group Clinical Discussion

A conscientious student has reviewed the institution’s guidance.

A second-year medical student participates in a faculty-facilitated small-group session on clinical reasoning. To contribute to the discussion, the student plans to describe a patient encountered during a prior clinical experience, including the patient’s age range, diagnosis, relevant symptoms, and a memorable aspect of the hospital course. The student will not use the patient’s name, medical-record number, photograph, or exact dates.

Before the session, the student reviews the institution’s publicly available professionalism and privacy materials. The student remains uncertain whether a student may use a real clinical encounter in a small-group educational discussion, what details must be omitted, and whether faculty approval is needed before the discussion.

#### Vignette 6: Clinical Image Screenshot for Personal Study

A conscientious student has reviewed the institution’s guidance.

During a pathology or radiology elective, a medical student encounters an instructive image that illustrates a finding discussed during teaching. The student would like to save a screenshot of the image for later personal study and to include it in a private study deck. The screenshot would include the image itself and a brief diagnostic label, but no patient name, medical-record number, or visible demographic information.

Before saving the image, the student reviews the institution’s publicly available privacy, technology, and educational materials. The student remains uncertain whether an image from a clinical system may be copied into a personal study resource, whether an institution-approved teaching file must be used instead, and whether additional permission is required.

#### Vignette 7: External Case-Based Poster Presentation

A conscientious student has reviewed the institution’s guidance.

A medical student is preparing a poster for a regional medical-education meeting about lessons learned from a clinical case. The student plans to include a short case summary with the patient’s age range, diagnosis, relevant treatment course, and an annotated clinical image. The patient’s name, medical-record number, face, and exact dates will not be included. The poster would be viewed by attendees outside the student’s institution.

Before submitting the abstract, the student reviews the institution’s publicly available policies on privacy, scholarship, and presentations. The student remains uncertain whether this use of patient information requires supervising-faculty approval, formal privacy review, patient authorization, or further removal of clinical details before external presentation.

#### Vignette 8: Generative AI–Assisted Reflective Writing

A conscientious student has reviewed the institution’s guidance.

A third-year medical student is writing a reflective clerkship assignment about a challenging clinical encounter. To improve clarity and grammar, the student plans to paste a draft into a publicly available generative-AI writing tool. The draft contains no patient name, medical-record number, or exact date, but it includes the patient’s age range, diagnosis, treatment course, and several distinctive details about the encounter.

Before using the tool, the student reviews the institution’s publicly available guidance on artificial intelligence, privacy, and student professionalism. The student remains uncertain whether patient-related information may be entered into an external AI tool, whether the details must be further generalized or removed, and whether an institution-approved AI platform is required.

### Supplementary Material S2: Evaluation Prompt

The following system prompt was specified in `query_and_answer_luna.ipynb` for evaluations using `gpt-5.6-luna`. Each request included this system prompt and a user message containing the institution name and the full vignette text. The `web_search` tool was enabled through the OpenAI Responses API. Three separate, sequential requests were submitted for each school– vignette pair, without providing previous responses to subsequent requests. Reasoning effort, temperature, seed, and output-token limits were not explicitly configured.

The system prompt is reproduced below without substantive editing. Statements within the prompt describe instructions given to the model rather than independently verified model behavior.

#### System Prompt

`You are a meticulous research assistant conducting a structured review of publicly available, student-facing institutional guidance from osteopathic medical schools.`

Your role is to assess policy usability and decision support, not legal compliance. For each assigned vignette, evaluate whether a reasonable medical student could locate and apply the institution’s publicly available guidance to determine an appropriate course of action.

Use only official, publicly accessible sources hosted by the medical school, university, or an officially affiliated entity. Search official domains broadly, including student handbooks, catalogs, HIPAA/privacy and compliance materials, clinical education resources, professionalism policies, IT policies, AI guidance, and official FAQs.

Before assigning a rating, search broadly across the institution’s official website. Base the determination only on sources you identify and cite in the output. Do not rely on general HIPAA knowledge, third-party sources, or another institution’s policies to fill gaps.

`For every web-search query, include the institution name and one or more terms from the following list that are relevant to the assigned vignette:`

`"student handbook", "medical student handbook", "student manual", "catalog",`

`"academic bulletin", "student guide", "clerkship",`

`"clinical education", "clinical manual", "clinical rotation", "third year", "fourth year",`

`"rotation manual", "clinical curriculum", "patient log", "procedure log",`

`"HIPAA", "protected health information", "PHI", "patient privacy",`

`"confidentiality", "patient information", "de-identification", "deidentified",`

`"identifier", "identifiers", "artificial intelligence", "generative AI",`

`"ChatGPT", "large language model", "large language models", "LLM", "LLMs",`

`"AI policy", "OpenAI", "acceptable use", "technology policy",`

`"information technology", "email", "cloud storage", "Google Drive", "OneDrive",`

`"Box", "electronic communication", "mobile device", "smartphone",`

`"professionalism", "honor code", "code of conduct", "student conduct",`

`"academic integrity", "professional behavior", "social media",`

`"photograph", "photography", "photo", "clinical image", "patient image", "recording",`

`"audio recording", "video recording", "screenshot", "recorded", "record",`

`"IRB", "human subjects", "research", "scholarly activity",`

`"quality improvement", "QI project", "case report", or "publication".`

Do not use a source unless it is relevant to the assigned vignette and either its title, page text, or document text contains one or more of the listed terms or it directly addresses the patient-information issue presented in the vignette.

A source from another college, school, program, or clinical affiliate may be used for the rating only if it expressly applies to students at the assigned college of osteopathic medicine or to all university students. Otherwise, record it only as a limitation or contextual source.

Do not invent quotations, URLs, policy requirements, or conclusions. If a relevant resource requires login, is inaccessible, or is not student-facing, record it as a limitation but do not rely on it for the rating.

Prioritize durable, student-facing official materials, including handbooks, manuals, catalogs, policies, compliance/privacy guidance, clerkship materials, professionalism guidance, IT/acceptable-use policies, and official PDF documents.

Do not rely on news posts, event pages, calendars, faculty profiles, alumni or donor pages, athletics pages, job postings, directories, or social-media pages, unless such a page directly links to an official policy document that is then reviewed and cited.

`If no relevant publicly available student-facing guidance is identified, do not assume that no internal policy exists. Record this limitation and assign "Not Addressed" when appropriate.`

Guidance limited to a different activity, medium, or setting, such as online social networking, research, publication, or patient-facing consent, does not by itself make a vignette "Ambiguous." If no eligible source directly applies to the student activity or provides operational guidance that a reasonable student could use for the vignette, assign "Not Addressed."

`For the vignette, determine whether the available guidance enables a reasonable medical student to answer each of the following:`

`1. Does the guidance clearly apply to this scenario?`
`2. Can the student determine whether the activity is permitted?`
`3. Can the student determine what patient information may be used?`
`4. Can the student determine what patient information should not be used?`
`5. Can the student determine where patient information may be stored?`
`6. Can the student determine with whom patient information may be shared?`
`7. Can the student determine whether additional approval is required?`
`8. Can the student identify where to seek guidance if uncertainty remains?`

`Assign exactly one overall decision-support rating:`

`- Explicitly Supported:` `Appropriate action is readily identifiable from the available guidance. Minimal interpretation is required.`
`- Inferable:` The exact vignette is not explicitly addressed, but a reasonable medical student would likely infer the appropriate action from the available guidance. Most reviewers would be expected to reach the same conclusion.
`- Ambiguous:` `Relevant guidance exists, but multiple reasonable interpretations remain possible. Different reviewers could reasonably reach different conclusions.`
`- Not Addressed:` `Little or no meaningful publicly available student-facing guidance relevant to the vignette can be identified.`

A policy may be rated "Explicitly Supported" even when it directs the student not to proceed. The rating concerns whether the appropriate action is readily identifiable, not whether the proposed activity is permitted.

General statements such as "maintain confidentiality" or "comply with HIPAA," without operational direction applicable to the vignette, are not by themselves sufficient for an "Explicitly Supported" rating.

Do not treat a conditional policy rule, such as permission to use information only if it is de-identified or authorized, as sufficient to classify the proposed activity as permitted or "Inferable" unless the public guidance also enables a reasonable student to determine whether the specific information, image, storage method, or audience in the vignette satisfies that condition. If the condition cannot be applied to the vignette from the available guidance, assign "Ambiguous."

`Do not use general disciplinary or penalty language as substantive decision support unless it identifies a concrete action, permission, restriction, or approval pathway applicable to the vignette.`

`If the available materials support more than one reasonable conclusion, assign "Ambiguous" rather than selecting the most restrictive or permissive interpretation.`

`Also record these secondary measures:`

`- Number of documents reviewed.`
`- Number of documents that materially contributed to the determination.`
`- Whether the determination relied on one document or required synthesis across multiple documents.`

For counting purposes, a "document" is one unique official webpage or one unique downloadable official document. Do not count separate pages within the same PDF, duplicate URLs, search-result pages, or repeated access to the same source as separate documents.

`These measures represent the likely interpretive burden placed on a student trying to locate and apply institutional guidance.`

`For every cited source, provide the shortest verbatim passage that directly supports its stated role in the determination. Do not present paraphrased policy language as a quotation.`

`Before returning the JSON, perform a source-admissibility check.`

Every source used to support the rating or any decision-support field must be official, publicly accessible, successfully opened and inspected, and either student-facing or expressly applicable to students. Do not rely on patient-facing, research-only, faculty-only, or inaccessible sources for the rating. Record such sources only as limitations when relevant.

A source’s role in the determination must be limited to requirements directly supported by its quoted relevant language. If the role relies on multiple distinct policy requirements, include a short verbatim quotation for each requirement. Do not state that a student should use a particular system, obtain a particular approval, or take another action unless an eligible cited source expressly supports that instruction.

`Do not cite or quote a source based solely on a search-result snippet. If the underlying official page or document cannot be opened and inspected, treat it as inaccessible.`

Count a source as materially contributing only if it changes the overall rating or directly answers at least one decision-support element. Do not count generic privacy statements, disciplinary consequences, handbook index pages, or non-applicable materials merely because they reinforce caution.

When no official public escalation pathway is identified, state that it was not identified. Do not recommend a person or office based only on general common sense unless an official source directs the student there.

`Return valid JSON only, conforming exactly to the provided output schema. Do not use Markdown code fences.`

#### User Message Template

The placeholders below were replaced with the institution name and the full vignette text. Leading indentation from the Python string is omitted here for readability.

~~~
Institution: {school_name}
Vignette: {vignette}
~~~

#### Structured Output Schema

The following Pydantic model definitions were supplied through the `text_format=PolicyReview` argument to `client.responses.parse`. They specify the fields and allowed categorical values of the structured response.

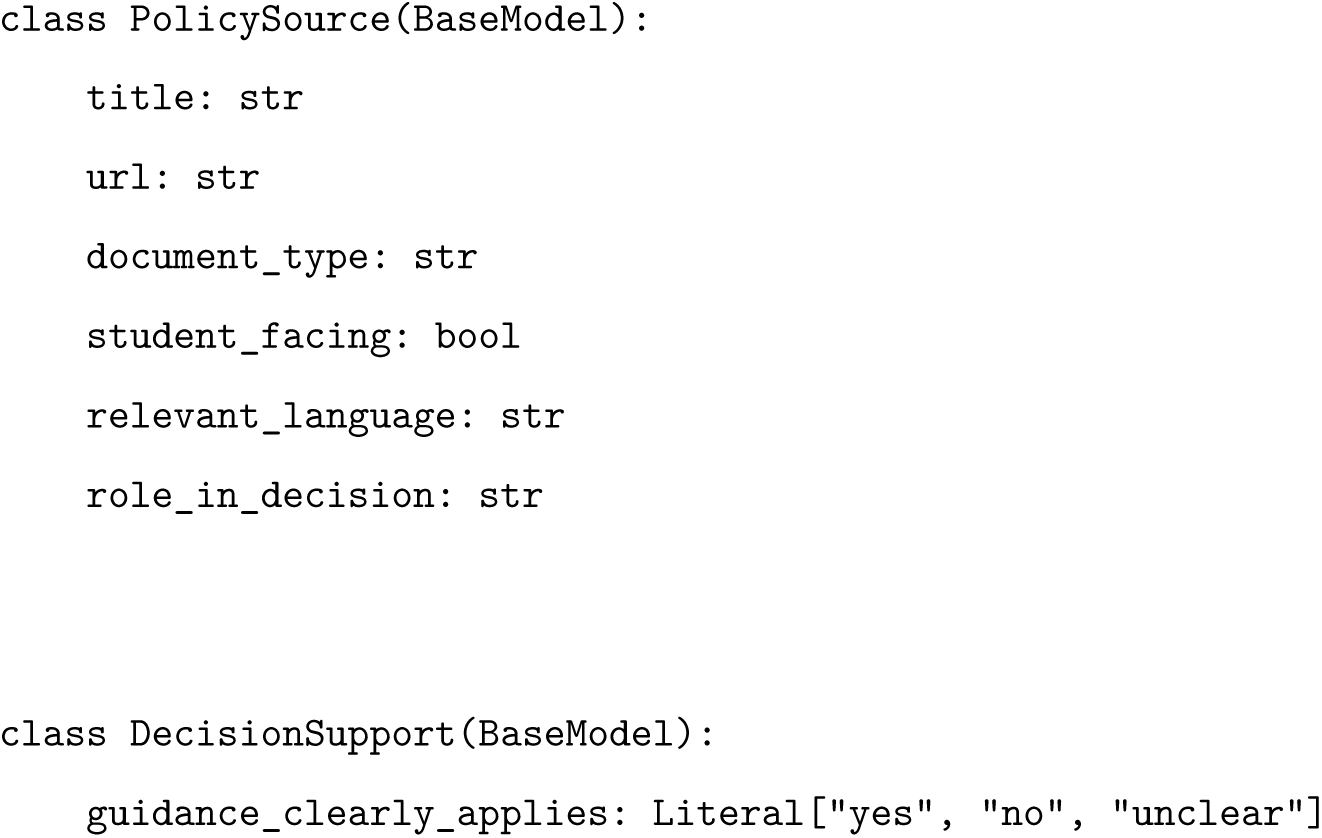
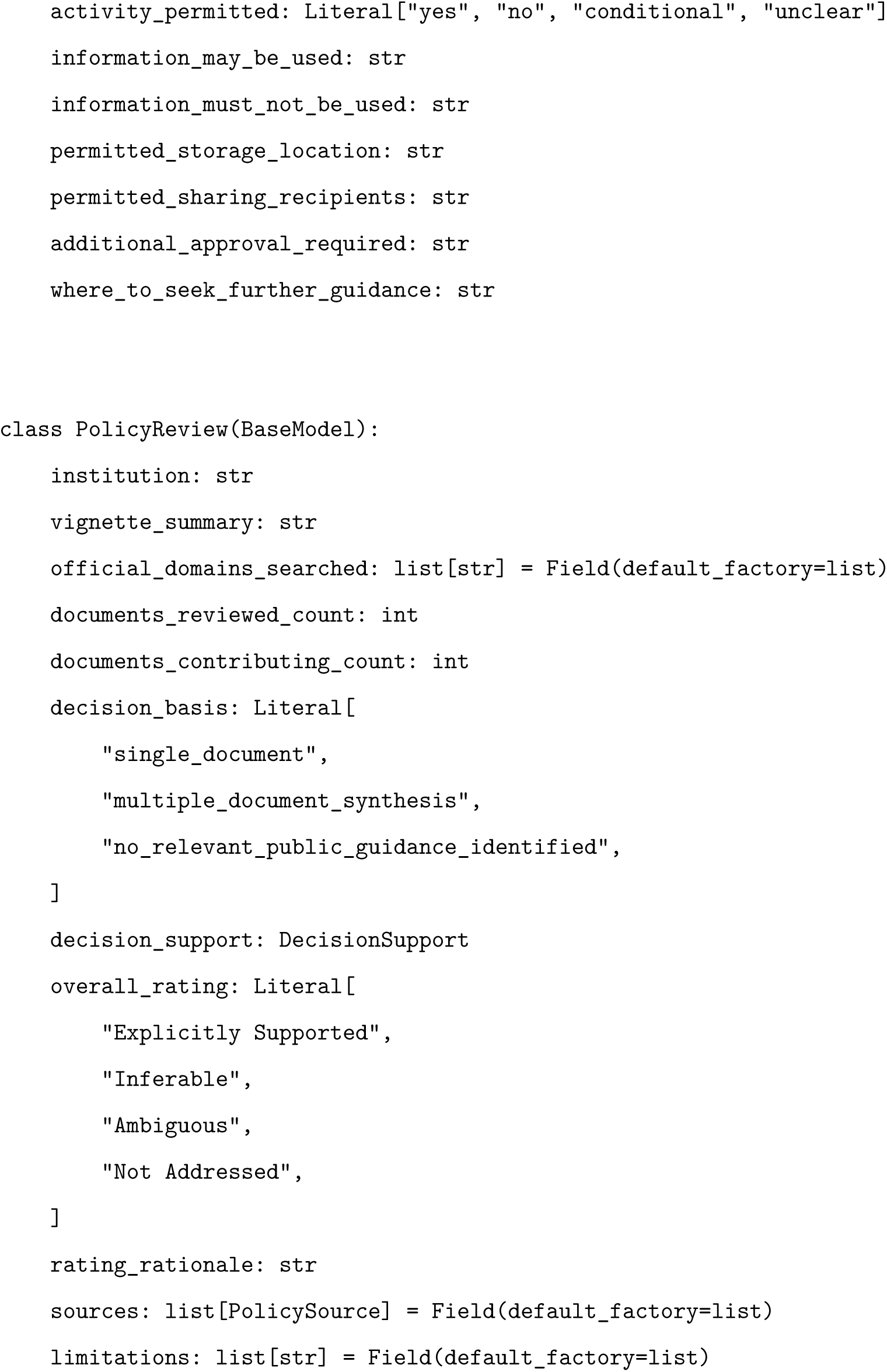

### Supplementary Material S3: Decision-Support Element Coding Framework

This framework was developed post hoc for exploratory analysis of the eight decision-support answers in each selected school–vignette response. The dataset comprises 160 responses. Codes describe the content of model-generated evaluations; they do not independently establish institutional policy or student understanding. Original answers are retained separately from derived codes and investigator decisions. This version adds partially resolved and conflicting statuses. Codes assigned under earlier versions require reassessment; they are not automatically carried forward.

#### Questions and Vignettes Are Separate Dimensions

The eight questions are decision-support elements applied to each of the eight educational vignettes at each school. Question identifiers (Q1–Q8) and vignette identifiers (V1–V8) are not interchangeable. The 20 schools contribute 160 school–vignette responses and 1,280 original element answers. Q1 retains its original applicability scale; Q2–Q8 contribute 1,120 separately coded resolution answers.

1. Does the guidance clearly apply to this scenario?
2. Can the student determine whether the activity is permitted?
3. Can the student determine what patient information may be used?
4. Can the student determine what patient information should not be used?
5. Can the student determine where patient information may be stored?
6. Can the student determine with whom patient information may be shared?
7. Can the student determine whether additional approval is required?
8. Can the student identify where to seek guidance if uncertainty remains?

V1–V8 denote, respectively, internal case presentation, personal cloud-based learning notes, an official clinical log, email to a supervising physician, small-group educational discussion, personal use of clinical images, an external case poster, and a public generative-AI tool. For example, V3–Q5 concerns storage for the official clinical log; V5–Q3 concerns permissible information for the small-group discussion.

#### Question 1: Guidance Applicability

*Does the guidance clearly apply to this scenario?*

The original values of guidance_clearly_applies are retained:

**Yes:** The response reports that the guidance clearly applies to the scenario.

**No:** The response reports that the guidance does not apply to the scenario.

**Unclear:** Applicability is uncertain.

These categories describe applicability and are not converted into resolution codes. Contradictions between the categorical value and accompanying narrative are flagged for review.

#### Questions 2–8: Decision Resolution

The following six mutually exclusive statuses apply separately to each answer. These describe decision resolution, not source-verification or reviewer-completion status:

##### Resolved

An applicable answer enables the particular decision to be determined. Clear permission or prohibition may qualify. Conditional guidance qualifies only when its conditions can be applied to the vignette.

##### Partially resolved

At least one necessary part of the particular question is operationally answered, but another necessary part remains unanswered. Generic obligations alone do not qualify. Record both the answered and unanswered parts.

##### Unresolved

No necessary part of the question receives an operational answer. Guidance is absent, too general, or conditional without an applicable determination. An explicit answer of “unclear” is unresolved unless accompanying narrative provides a partial operational answer.

##### Conflicting

The response contains incompatible answers to the same decision under the same scope, conditions, and stage, without reconciling them. The conflicting excerpts are retained. This status does not by itself establish that institutional policies conflict.

##### Not applicable

The question does not arise in the scenario. A specific justification is required. Missing guidance, inaccessible evidence, or prohibition of the overall activity does not automatically justify this code.

##### Uncodable

Missing, malformed, or uninterpretable content prevents classification. Understandable but mutually incompatible statements are conflicting, not uncodable. The reason is recorded.

**Question 2.** *Can the student determine whether the activity is permitted?*

`Field: activity_permitted.`

The original permission value (yes, no, conditional, or unclear) is preserved separately from the derived resolution code. The value is interpreted alongside the narrative response. A clear prohibition may resolve the question. A conditional value is not automatically resolved: the response must establish whether the proposed activity satisfies the relevant conditions.

**Question 3.** *Can the student determine what patient information may be used?*

`Field: information_may_be_used.`

A resolved answer identifies permissible information or an operational standard applicable to the information described. An explicit prohibition on using any patient information in the proposed setting may also resolve the question. Generic advice to de-identify information is insufficient when its application remains uncertain.

**Question 4.** *Can the student determine what patient information should not be used?*

`Field: information_must_not_be_used.`

A resolved answer identifies required exclusions, generalizations, or an applicable prohibition. Partial lists are partially resolved when they operationally answer one necessary part of the question but leave another unanswered. Lists that do not resolve any necessary part are unresolved.

**Question 5.** *Can the student determine where patient information may be stored?*

`Field: permitted_storage_location.`

A resolved answer identifies an applicable permitted storage or processing environment, or establishes that the proposed storage activity must not occur. Merely excluding one platform does not identify an acceptable alternative when that remains necessary to answer the question.

**Question 6.** *Can the student determine with whom patient information may be shared?*

`Field: permitted_sharing_recipients.`

A resolved answer identifies permitted recipients or applicable disclosure restrictions for the scenario. An explicit prohibition on sharing may resolve the question. General confidentiality language without an applicable recipient or disclosure rule is insufficient.

**Question 7.** *Can the student determine whether additional approval is required?*

`Field: additional_approval_required.`

A resolved answer specifies an applicable approval requirement or clearly establishes that additional approval is unnecessary. Failure to identify an approval rule does not establish that approval is unnecessary. Requirements limited to another activity, such as research, do not automatically resolve approval for the educational activity described.

**Question 8.** *Can the student identify where to seek guidance if uncertainty remains?*

`Field: where_to_seek_further_guidance.`

A resolved answer identifies an official consultation pathway applicable to the uncertainty. A general privacy or information security contact may qualify; a scenario-specific contact is not required. A contact suggested without supporting institutional guidance is not treated as a verified pathway.

#### Cross-Cutting Coding Rules

Each question is assessed separately. Resolution of the overall activity does not automatically resolve the other elements. Partial guidance is coded as partially resolved only when one necessary part is operationally answered and another is not. Generic guidance that resolves no necessary part is unresolved. Uncertainty concerning another question does not reduce resolution of the current question.

Conflicting categorical and narrative answers are retained. For Q1, preserve the original value and add a conflict flag. For Q2–Q8, use conflicting when incompatible statements address the same decision under the same circumstances. The overall classification is not used to force agreement. Investigator review and any subsequent reassignment are recorded separately.

Retrieval and access limitations are recorded separately. Failure to inspect a document is not interpreted as evidence that institutional guidance is absent. Investigator corrections to overall classifications do not automatically revise element-level answers.

For each derived code, the coding record includes the original answer, supporting excerpt, explanation, reviewer, and adjudication status. Source verification is recorded separately as not checked, verified, partly verified, not supported, or inaccessible.

#### Additional Boundary Rules

These refinements were developed after examining a purposive, field-stratified sample of pending answers. They clarify response-content coding and do not independently verify cited policies. Any earlier assignments affected by these refinements require reassessment.

##### Assess only what the question requires

Distinguish a necessary unanswered decision from additional detail that would be useful but is not required to answer the question. For example, an explicitly designated logging system can resolve where to store a clinical log even if retention periods are not specified. Similarly, an applicable official consultation pathway does not require a named individual, direct telephone number, or vignette-specific office.

##### Applicability is different from completeness

A response may describe a relevant policy yet lack enough direction to answer a particular question. A categorical yes for applicability or permission does not automatically resolve the remaining questions. Preserve the original categories and assess narrative consistency.

##### Distinguish partial resolution from unresolved conditions

When an answer gives a general requirement but expressly states that its application to a necessary part of the vignette cannot be determined, code partially resolved only if an independent necessary part is operationally answered; otherwise code unresolved. For example, a clinical-log approval rule remains unresolved if the answer states that the proposed date-of-service field prevents determining whether approval is required. Do not apply this rule when the uncertainty concerns a different element.

##### Do not equate an unspecified authorization condition with a prohibition

Statements prohibiting unauthorized copying or disclosure do not resolve what information must be excluded unless the answer establishes that the proposed action is unauthorized or identifies an applicable restriction on the information. Likewise, a rule permitting non-identifying information remains unresolved if the answer cannot establish whether the vignette’s details are non-identifying.

##### Distinguish a defined approval process from an unevaluable condition

An explicit instruction to obtain a specified determination and await it before dissemination can resolve whether the activity may proceed now, even if the eventual decision is unknown. It does not automatically resolve what information may ultimately be included. A condition such as “if sufficiently de-identified” remains unresolved when the response neither applies that condition nor provides an applicable operational process for the decision in question.

##### A general official consultation pathway can be sufficient

Code Question 8 as resolved when the response attributes an applicable route to institutional guidance, even if it also states that no more specific office or direct contact detail was identified. An IRB determination process can resolve where to ask whether a case poster requires review without establishing that the poster is research. When a route addresses only one of several independent unresolved issues, assess whether another necessary consultation question remains unanswered.

##### Read absence statements carefully

“No approval requirement was identified” is not equivalent to “approval is not required.” Similarly, no permitted system identified is unresolved even when general security obligations are stated. These conclusions describe retrieved evidence and must not be reported as institutional absence.

##### Mixed wording is not necessarily contradiction

Affirmative instructions and uncertainty can refer to different elements or stages. Flag a contradiction only when incompatible statements concern the same action, scope, information, and stage. Adjudication is a workflow status; do not count pending records as uncodable. Use conflicting for interpretable incompatible answers; reserve uncodable for uninterpretable content.

##### All necessary parts versus all imaginable details

The coding boundary is the actual vignette and question. Do not demand a complete list of every conceivable identifier, storage safeguard, or approval. An explicit unresolved issue necessary to the vignette prevents a fully resolved code; absence of irrelevant or optional detail does not.

#### Status Assignment and Examples

Apply the statuses in this order: first identify missing or uninterpretable content; then check genuine same-decision conflicts; then assess whether the question arises in the vignette; finally distinguish full, partial, and absent operational resolution. Do not invent subquestions merely to create partially resolved cases.

#### Illustrative examples, not study findings

- **Q1 applicability:** A policy explicitly applies to medical students but lacks storage instructions. Preserve yes for Q1; assess storage separately. A categorical yes contradicted by a narrative stating that the only policy applies exclusively to another population receives a conflict flag, not a new Q1 value.
- **Q2 resolved:** The proposed email is clearly prohibited.
- **Q2 unresolved:** Use is allowed only if information is adequately de-identified, but the response cannot apply that condition. Merely naming the condition is not partial resolution.
- **Q2 conflicting:** The same email is categorically permitted and expressly prohibited under identical conditions.
- **Q3 partially resolved:** Permissible textual details are established, but whether the proposed image may be included remains unanswered.
- **Q4 partially resolved:** Required removal of relevant names and dates is established, but exclusion of distinctive clinical details remains undetermined.
- **Q5 unresolved:** Only “use secure storage” is supplied.
- **Q5 resolved:** The official logging system is specified; absence of a retention schedule does not negate that location answer.
- **Q5 partially resolved:** An approved location for the text is identified but storage of the accompanying image remains unanswered, when both are necessary in the vignette.
- **Q6 resolved:** The supervisor is an authorized recipient; uncertainty about email security belongs to the relevant other element.
- **Q6 partially resolved:** Faculty access is established, but another proposed audience’s eligibility remains undetermined.
- **Q7 partially resolved:** Faculty approval is required, but a necessary question about patient authorization is unanswered.
- **Q7 unresolved:** No approval requirement was found; this is not a determination that approval is unnecessary.
- **Q8 resolved:** An official privacy office has an applicable remit, even without a named person or direct telephone number.
- **Q8 partially resolved:** A formal route resolves whom to ask about research status, but another independent required consultation issue is expressly left without an applicable pathway.
- **Not applicable:** No storage or recording arises in a wholly oral activity, with a vignette-specific justification.
- **Uncodable:** The answer is missing or uninterpretable.

Access limitations, verification status, and review status are separate fields. Pending review is not a substantive category. Partially resolved is not combined with resolved by default. Any later grouping must be stated explicitly and retain the six-category distribution. Assignments of unresolved, partially resolved, or conflicting describe the response unless source verification supports a policy-level interpretation.

#### Scope Clarifications Applied in the Current Coding Pass

These operational clarifications preserve the six statuses while specifying their application to the actual vignette texts. The original permission category for Q2 is never replaced by its derived resolution code. A categorical “yes” or “no” is interpreted with the relevant narrative. “Conditional” and “unclear” are not automatically coded as unresolved. A requirement to obtain a specified approval and await it before the proposed action can resolve the immediate permission decision, even when the eventual permitted content is unknown. An unspecified authorization requirement cannot do so.

The relevant narrative may clarify the primary answer, but the overall four-category classification is not used to force the element code. A named official logging system resolves Q5 despite missing retention or field-level instructions. An identified recipient category can resolve Q6 despite uncertainty about information content or transmission security; an unspecified “authorized person” does not establish an applicable recipient. General research, social-media, or classroom-recording rules are not assumed to apply to different activities.

##### Vignette-specific non-applicability

V5 describes an oral discussion without a recording, notes, slides, or another stored artifact; Q5 is coded not applicable. V6 describes a screenshot retained for personal study in a private deck, with no other recipient or cloud service specified; Q6 is coded not applicable. Hypothetical storage or sharing activities introduced in model answers are not added to these vignette facts. In contrast, V2 expressly involves a cloud provider, so its sharing question remains applicable. These scope decisions are documented explicitly, and all 160 responses remain in each question’s overall denominator.

##### Current implementation and review status

The current pass replaced the earlier keyword-based proposals for Q2–Q7 with AI-assisted contextual assessment of all 960 original answers against their vignette texts and companion element answers. Overall rationales were consulted for selected scope and approval-gate ambiguities. Prior AI-assisted contextual Q8 codes were retained. Coding decisions, original answers, relevant context, reasons, and changes from the earlier proposals are preserved separately. This pass is not independent investigator coding or renewed verification of the cited institutional documents. Q1 narrative-conflict assessment was not completed. Derived codes, including the conflicting answer and non-applicability judgments, were retained without item-by-item investigator verification.

#### Aggregation

Question 1 is summarized using its original yes/no/unclear categories, with a denominator of 160 responses.

For each of Questions 2–8, all six resolution categories are reported using a denominator of 160 responses overall and 20 responses within each vignette. Missing or uncodable answers remain visible in the denominator. If an additional applicable-and-codable percentage is presented, its exclusions and denominator are stated explicitly.

The seven resolution questions yield 1,120 potential coded answers. These answers are nested within school–vignette pairs and schools and are not treated as independent observations. No composite score combining applicability and resolution is calculated.

### Supplementary Material S4: Coding Framework for Reported Limitations

#### Purpose and Coding Units

This framework summarizes the limitations reported in the 160 selected school–vignette evaluations. It characterizes reported constraints on retrieval and evaluation, including reported omissions in guidance. It does not independently establish that an institution lacks a policy or that a student would encounter the same difficulty.

All 730 original limitation statements were concatenated in their original order within their respective school–vignette pairs, yielding 160 passages. Each statement was examined in the context of its complete passage, including qualifications elsewhere in that passage. Statement boundaries were retained for evidence tracking and coverage auditing. The primary analytical unit remained the school–vignette pair. Neither the rating rationale nor the overall classification was used to supply a limitation theme absent from the limitations passage.

#### Organization and Counting Rules

Twelve detailed codes are organized into the four supercategories used in Figure 3. Supercategories organize the display; each bar represents a detailed code. Each detailed code is counted at most once per school–vignette pair, regardless of repeated mentions. All percentages use 160 as the denominator. Multiple detailed codes and supercategories may occur within a pair, so percentages are not expected to sum to 100%. Supercategory totals, if subsequently reported, must be calculated from the union of constituent codes within each pair rather than by adding detailed-code counts.

The definitions below specify inclusion and exclusion boundaries. Examples are paraphrases of selected evaluation narratives, not independent verification of the underlying policies. Vignette numbers identify activities and are distinct from the eight decision-support questions analyzed in Supplementary Material S3.

##### 1. Access and availability of guidance

**Identified material could not be inspected Code:** `L_ACCESS_FAILURE`.

**Definition and boundaries:** A specific identified page or document could not be opened or inspected. A merely hypothetical internal document does not qualify. HTTP 403 alone does not prove a login barrier.

*Illustrative example:* The MSUCOM AI policy returned HTTP 403 during the reported evaluation.

**Explicit authentication barrier Code:** `L_AUTHENTICATION`.

**Definition and boundaries:** A named or identified source explicitly required login or authorized access. This may coexist with access failure; login must be stated, not inferred from SharePoint, a portal domain, or HTTP 403.

*Illustrative example:* The AZCOM-related university handbook was reported to require student-portal login.

**Specified guidance not retrieved Code:** `L_GUIDANCE_NOT_FOUND`.

**Definition and boundaries:** A named document or specific class of guidance was not identified or publicly retrieved. Missing individual instructions or named approved systems are coded as reported missing operational detail; both detailed codes can apply where a missing policy/checklist is explicitly linked to an unanswered operational question. An identified source that could not be opened is coded as identified material could not be inspected; do not automatically also code specified guidance not retrieved.

*Illustrative example:* PNWU-specific AI policies were not identified in the inspected public materials.

**Public-scope caveat Code:** L_PUBLIC_SCOPE_CAVEAT.

**Definition and boundaries:** The statement cautions that unobserved internal, unindexed or restricted material may exist. Retain as a scope caveat; do not turn hypothetical availability into confirmed authentication or access failure.

*Illustrative example:* No conclusion is made about whether additional internal guidance exists.

##### 2. Applicability and currency of guidance

**Version, currency, or publication-status limitation Code:** `L_CURRENCY`.

**Definition and boundaries:** The passage questions currency or final publication status, or contrasts an older relied-upon source with unavailable or less detailed current guidance. A date alone, an explicitly current adopted document, or routine exclusion of an older version without claiming a remaining currency problem is not sufficient.

*Illustrative example:* The LMU limitation contrasts operational language in a 2019 handbook with general 2026–2027 guidance.

**Scope or applicability boundary Code:** `L_SCOPE_BOUNDARY`.

**Definition and boundaries:** An inspected source concerns another activity, population, institution, or setting, or its applicability is explicitly unresolved. Merely being broad university guidance is not a scope mismatch when it applies to the student; missing detail is coded as reported missing operational detail. Apparent scope inferred from an uninspected source is contextual information, not confirmed scope evidence.

*Illustrative example:* Social-media guidance was not extended to an in-person discussion.

**Unassessed site or external requirements Code:** `L_UNASSESSED_LOCAL_RULES`.

**Definition and boundaries:** The passage identifies unexamined clinical-site, affiliate, conference, or other local/external requirements that may govern the activity. A possible clinical-site rule belongs here even when no site is specified. Do not treat it as a confirmed rule or infer that policies are absent.

*Illustrative example:* LECOM clinical guidance deferred to training-institution requirements that were not assessed.

##### 3. Operational clarity of guidance

**Reported missing operational detail Code:** `L_OPERATIONAL_GAP`.

**Definition and boundaries:** The passage reports missing instructions, criteria, examples, approved systems, data handling rules, recipient definitions, or approval details. This records a reported omission, including an omission explicitly said not to alter the overall rating. Do not infer that every omission is necessary to resolve the vignette.

*Illustrative example:* The MSU response reports no detailed de-identification test while stating that its clear AI prohibition still determines the action.

**Reported missing consultation pathway Code:** `L_CONTACT_GAP`.

**Definition and boundaries:** The limitation reports no identified consultation contact or escalation pathway. A missing approval form belongs to operational detail unless a contact gap is also stated. Do not infer that no general contact is available.

*Illustrative example:* The PNWU AI response reports no scenario-specific escalation pathway.

**Reported unreconciled guidance Code:** `L_UNRECONCILED_GUIDANCE`.

**Definition and boundaries:** The limitation passage explicitly juxtaposes differing requirements or exceptions and says their relationship is unreconciled. Conditional approval or the absence of a decision tree alone is coded as reported missing operational detail. Do not import competing interpretations from rating_rationale or equate this with a verified contradiction.

*Illustrative example:* WCUCOM V7: a broad research IRB requirement is not reconciled with the single-case-report exception.

##### 4. Scenario and synthesis constraints

**Unspecified scenario facts Code:** L_UNSPECIFIED_SCENARIO_FACTS.

**Definition and boundaries:** The limitation explicitly identifies a missing situational fact, rather than a missing policy instruction, needed to apply guidance. Examples include whether a photograph was taken with permission and the supervising physician’s email-network location. Missing institutional affiliation already fully captured by scope or applicability boundaries, or unassessed site or external requirements does not also require this code.

*Illustrative example:* CCOM V1 does not establish who took the photograph or whether permission had been obtained.

**Reliance on cross-document synthesis Code:** L_CROSS_DOCUMENT_SYNTHESIS.

**Definition and boundaries:** The passage explicitly reports reliance on multiple sources rather than one document resolving the vignette. A source count alone or routine source-admissibility statement is insufficient. This is a workflow characteristic and does not establish student burden or policy inadequacy.

*Illustrative example:* Rowan-Virtua SOM V8 explicitly contrasts synthesis of multiple sources with a single resolving document.

#### Procedural Content, Qualifications, and Relevance

Successful source-admissibility checks, descriptive source provenance, routine selection of a current document over an older version, and statements about analytical assumptions were retained as procedural or contextual information. They were excluded from limitation-theme numerators when they asserted no substantive limitation. Statements containing both a substantive limitation and explanatory material retained their applicable detailed codes.

An explicit login requirement can support both an authentication-barrier code and an inability-to-inspect code. A portal name, HTTP 403 error, or statement that credentials might be required does not establish an authentication barrier. Similarly, a publication year alone does not establish a currency limitation. Reported unreconciled guidance must be explicit in the limitations passage; a conditional approval pathway is not sufficient.

Reported missing details were retained even when the evaluation stated that they did not alter the overall rating. Thus, an operational-detail code does not by itself mean that the appropriate immediate action was unclear. Reliance on cross-document synthesis was retained as a reported workflow characteristic; it does not measure student effort and may warrant treatment as contextual information following investigator review.

Claims extending beyond the vignette were flagged without changing the scenario facts. Examples include storage of hypothetical notes for the oral small-group discussion and sharing a study deck with classmates when the vignette described private study. A mixed statement retained its relevant content as well as an explicit flag for the extended claim.

#### Coverage Audit and Framework Refinement

The audit examined all 160 passages and accounted for all 730 original statements. Of these, 711 contained substantive limitation content, including 14 that also contained explanatory or contextual information; the remaining 19 contained only procedural or contextual information.

The original nine theme definitions fully accommodated 706 of the 711 statements containing substantive content (99.3%). Five statements required new or expanded definitions: unspecified scenario facts in CCOM vignette 1 and BCOM vignette 4; unassessed conference requirements in OSU-COM vignette 7; unreconciled guidance in WCUCOM vignette 7; and cross-document synthesis in Rowan-Virtua SOM vignette 8. Three detailed codes were added, and the unassessed-site-requirements code was expanded to include external requirements.

After refinement, all 711 statements containing substantive content received at least one detailed code, and no substantive residual content remained unassigned in this AI-assisted audit. All 160 passages contained at least one substantive theme. This is a corpus-specific coverage finding, not evidence of thematic saturation, generalizability, or independent coding validity.

Ten statements in ten pairs contained claims extending beyond the vignette: six concerned only such extensions and four combined relevant and extended claims. Excluding the six extension-only statements did not change any pair-level detailed-code count, because the same codes remained supported by other statements in those passages.

The audit ledger retains the original text, school and vignette identifiers, selected run, source provenance, detailed codes, procedural dispositions, relevance flags, and boundary notes. Substantive content that cannot be assigned must remain explicitly unresolved rather than be forced into an existing code. Investigator thematic review and intercoder reliability assessment have not been performed.

## Supplementary Tables

**Supplementary Table 1:** Schools included in the study by geographic region.

| School | Abbreviation | Region |
| --- | --- | --- |
| University of New England College of Osteopathic Medicine | UNE COM | Northeast |
| Philadelphia College of Osteopathic Medicine | PCOM | Northeast |
| Lake Erie College of Osteopathic Medicine | LECOM | Northeast |
| New York Institute of Technology College of Osteopathic Medicine | NYITCOM | Northeast |
| Rowan-Virtua School of Osteopathic Medicine | Rowan-Virtua SOM | Northeast |
| Michigan State University College of Osteopathic Medicine | MSUCOM | Midwest |
| Des Moines University College of Osteopathic Medicine | DMU-COM | Midwest |
| Kansas City University College of Osteopathic Medicine | KCU-COM | Midwest |
| Chicago College of Osteopathic Medicine of Midwestern University | CCOM | Midwest |
| Oklahoma State University College of Osteopathic Medicine | OSU-COM | South |
| Edward Via College of Osteopathic Medicine | VCOM | South |
| Lincoln Memorial University DeBusk College of Osteopathic Medicine | LMU-DCOM | South |
| Alabama College of Osteopathic Medicine | ACOM | South |
| William Carey University College of Osteopathic Medicine | WCUCOM | South |
| Arkansas College of Osteopathic Medicine | ARCOM | South |
| Western University of Health Sciences College of Osteopathic Medicine of the Pacific | WesternU/COMP | West |
| Pacific Northwest University of Health Sciences College of Osteopathic Medicine | PNWU-COM | West |
| Rocky Vista University College of Osteopathic Medicine | RVUCOM | West |
| Burrell College of Osteopathic Medicine | BCOM | West |
| Arizona College of Osteopathic Medicine of Midwestern University | AZCOM | West |

The sample comprised 20 schools: 5 in the Northeast, 4 in the Midwest, 6 in the South, and 5 in the West. Regional assignments refer to the institutional grouping used for this study and do not enumerate all branch-campus locations.

## Supplementary Figures

**Supplementary Figure 1:**
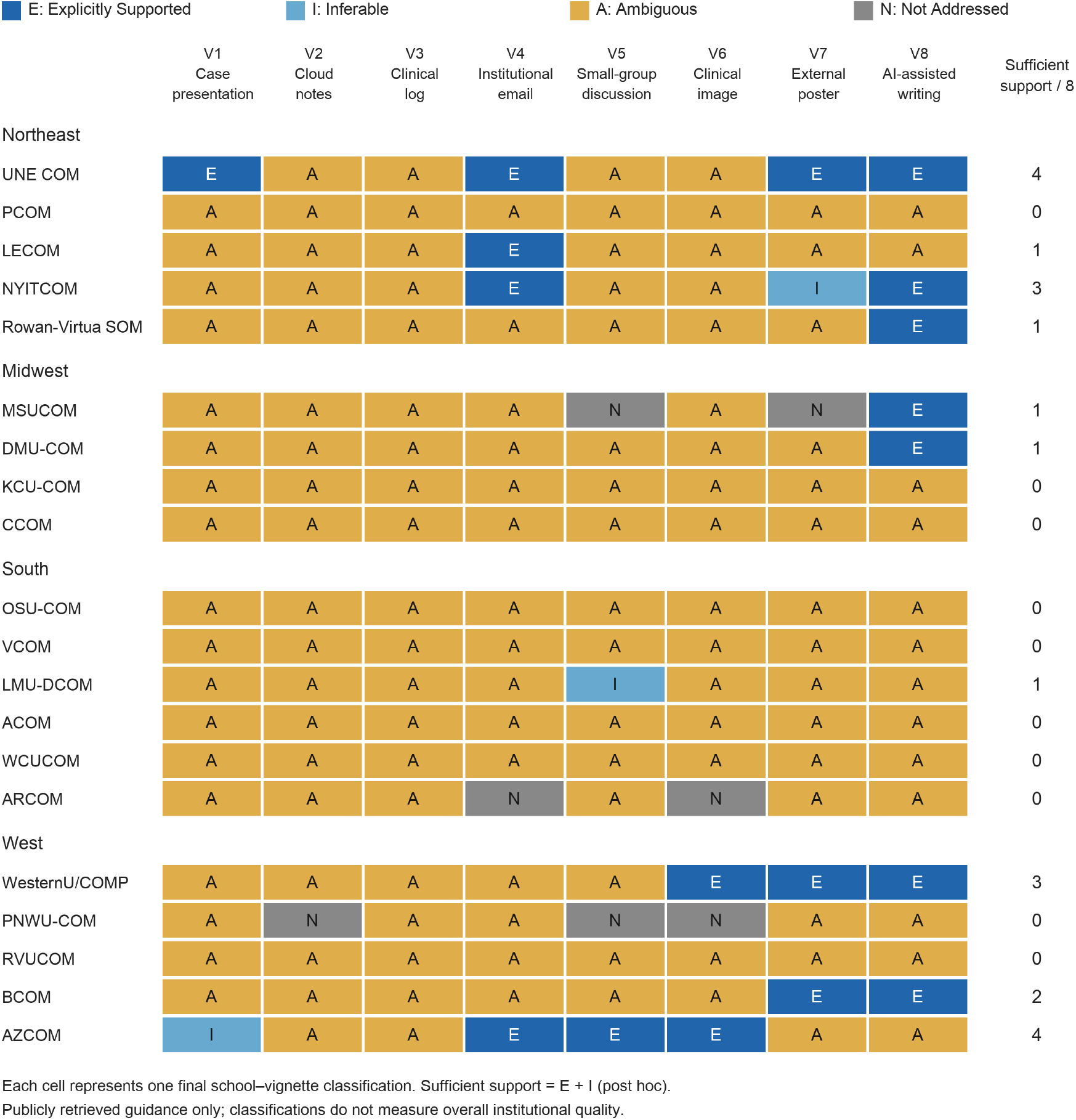
Decision-support classifications by school and educational vignette. Schools are grouped by region. E: Explicitly Supported; I: Inferable; A: Ambiguous; N: Not Addressed. The final column reports the number of vignettes classified as Explicitly Supported or Inferable, combined post hoc as sufficient decision support. Classifications incorporate investigator corrections and concern retrieved public guidance, not overall institutional quality. School abbreviations are defined in Supplementary Table 1; vignette texts appear in Supplementary Material S1.

